# Long-term Trajectories of Depressive Symptoms in Deployed Military Personnel: A 10-year prospective study

**DOI:** 10.1101/2023.08.14.23294068

**Authors:** Xandra Plas, Bastiaan Bruinsma, Caspar J. van Lissa, Eric Vermetten, Remko van Lutterveld, Elbert Geuze

**Affiliations:** Brain Research and Innovation Centre, Ministry of Defence, Utrecht, the Netherlands; Department of Psychiatry, University Medical Centre Utrecht Brain Centre, Utrecht, the Netherlands; Department of Methodology and Statistics, Tilburg University, Tilburg, the Netherlands; Department of Psychiatry, Leiden University Medical Centre, Leiden, the Netherlands

**Keywords:** trajectories, long-term depression, PTSD, miliary personnel

## Abstract

**Background:** Military missions are associated with an increased risk of depression. Understanding the long-term development of depressive symptoms after deployment is important to improve decision-making regarding deployment and mental health policies in the military. Therefore, this study aims to investigate trajectories of depressive symptoms from pre- to post-deployment and assess the role of specific factors, such as demographics, early life trauma, posttraumatic stress disorder (PTSD) symptoms and deployment stressors, in the Dutch army.

**Methods:** The study includes a cohort of 1032 military men and women deployed to Afghanistan between 2005 and 2008. From pre-to 10 years post-deployment (across 6 distinct time points), depressive and PTSD symptoms were assessed using the Symptom CheckList-90 (SCL-90) and the Self-Rating Inventory for PTSD (SRIP) respectively. Demographics, early trauma, and deployment experiences were collected at baseline and after deployment, respectively. Latent Class Growth Analysis was used to explore heterogeneity in developmental trajectories of depressive symptoms over time.

**Results:** The study identified four trajectories for depressive symptoms: resilient (65%), intermediate-stable (20%), symptomatic-chronic (9%), and late-onset-increasing (6%). The late-onset-increasing group had the highest proportion of individuals younger than 21 years. In addition, the resilient group was less likely to have experienced deployment stressors and the symptomatic-chronic group reported more early life traumas compared to the other groups. For individuals classified in trajectories with higher levels of depressive symptoms, PTSD symptoms were higher at all time points.

**Conclusions:** Multiple trajectories for depressive symptoms were identified in a military sample up to 10 years after deployment. These trajectories were associated with age, early trauma, deployment stressors and PTSD symptoms. The majority of the sample fell within the resilient trajectory, supporting the notion that deployed military personnel possess a high level of resilience. Overall, these findings provide valuable insights and a foundation for further research in this area.

## Introduction

Between 2005 and 2008 the Dutch armed forces participated in the International Security Assistance Force (ISAF) of NATO with either the Provincial Reconstructions Teams or with the Task Force Uruzgan. It is known that military missions such as ISAF, can have an impact on mental health, including depressive symptoms [2]–[4]. These adverse effects can present themselves not just in the short-term, but also long-term. Previous research from our group found long-term significant increases of depressive symptoms [5], even up to 10 years post-deployment [6]. Therefore, it is important to gain insight in the long-term development of depression in deployed military personnel to improve future deployment decisions or mental health care policies in the military. Additionally, these data give us the opportunity to learn more about the long-term development of depressive symptoms after a life stressor in general.

While the occurrence of depression in the military has been studied extensively, little attention has been paid to the long-term development (trajectories) of depression after deployment. Studies investigating trajectories of depressive symptoms mainly focused on civilian rather than military samples [7]–[10]. Musliner and colleagues [11] conducted a systematic review on the heterogeneity in long-term development of depressive symptoms and found that most of these studies identified three or four different symptom trajectories and sampled the general population. Only a small portion of the studies investigated depression trajectories in a military sample. Karstoft and colleagues [12] studied the trajectories of depressive symptoms from pre-to post-deployment over a period of 6.5 years in Danish soldiers. They found three trajectories: 1) low-stable (86.5%), 2) medium-fluctuating (4.0%) and 3) low-increasing (9.4%). Two other military sample studies by Sampson and colleagues [13], [14] found four trajectories of depressive symptoms in the United States army over 4 years: 1) resistant (55.8-61.5%), 2) increasing (12.8-13.2%), 3) decreasing (16.1-18.7%) and 4) chronic (9.2-12.7-%). As mentioned above, soldiers can experience depressive symptoms up to 10 years after deployment, but previous studies only covered periods that were significantly shorter. This study aims to broaden the knowledge of long-term development of depressive symptoms in military personnel by including a 10-year follow-up measurement.

A valuable tool for investigating heterogeneity in development is Growth Curve Modelling (GCM), a statistical method that analyses inter-individual variability in intra-individual patterns over time [15]. One specific type of GCM is Latent Class Growth Analysis (LCGA), which considers unobserved heterogeneity (different groups) over time within a larger population [16]–[18]. In other words, LCGA can examine the growth and shape of the development of depressive symptoms over time and assess how individuals in the population group together based on their symptom patterns.

Multiple factors might be associated with the development of depressive symptoms. For example, deployment, and particularly deployment with combat exposure, can increase the risk of developing depression [4]. Additionally, being female, poor education, prior trauma, and rank were found to be associated with depression or depression trajectories [11], [12], [19]. Knowledge about the factors associated with depression can help identify soldiers at risk for depression post-deployment. Also, depression often co-occurs with posttraumatic stress disorder (PTSD), a psychiatric disorder that can occur following exposure to traumatic experiences, which is common in the military context [20]–[23]. The comorbidity of PTSD and depression is more prevalent among military samples compared to civilian samples [24]. Here, we aim to expand the understanding of the factors linked to depression, as well as the co-occurrence of depressive and PTSD symptoms.

The aim of the present study is to investigate the development of depressive symptoms over a period of 10 years post-deployment in military personnel and assess the role of specific factors. To achieve this goal, GCM will be performed on data from the Prospective Research in Stress-Related Military Operations (PRISMO) cohort [25]. The PRISMO study was initiated in 2005 by the Dutch Ministry of Defence with the aim of studying the biological and psychological factors related to mental health both longitudinally and prospectively. 1032 soldiers participated in the PRISMO study in which data was collected at multiple time points over a period of 10 years from pre-to post-deployment. The PRISMO cohort has yielded several scientific insights including the impact of sleep [26] and biological factors [27]–[29] on fatigue, PTSD, and depression. However, there remains an unexplored area within the PRISMO cohort concerning long-term depression and its associated risk factors. Based on previous literature [11], we hypothesize that three or four trajectories of depression can be identified in deployed military personnel. Given that deployment increases the risk for depression [4] and considering the common co-occurrence of PTSD and depression [24], we additionally hypothesize that individuals following more resilient trajectories will be less likely to experience deployment stressors and exhibit reduced levels of PTSD symptoms.

## Methods

### Design and participants

Full details of the PRISMO cohort and measurement protocol can be found in the PRISMO protocol paper [25]. In brief, a total of 1032 military men and women participated in this prospective observational cohort study. All participants were deployed to Afghanistan between March 2005 and September 2008 on behalf of the ISAF mission. Data was collected over a period of 10 years (Figure 1). Approximately 1 month before deployment the baseline (T0) measurement was carried out at the army base. Two follow-up measurements were completed 1 month (T1) and 6 months (T2) after deployment at the army base. The 1-year (T3), 2-year (T4), and 5-year (T5) follow-up measurements were completed at home. Subsequently, the 10-year (T6) follow-up was completed at the research facility of the Military Mental Healthcare. Participants completed multiple questionnaires with paper-and-pencil (T0-T4, T6) or online (T5). Participants provided written informed consent prior to the study and all procedures were approved by the Institutional Review Board of the University Medical Centre Utrecht (Utrecht, The Netherlands).

### Measures

#### Depression

For T0-T4 and T6, depressive symptoms were assessed with the depression subscale of the Dutch Symptom CheckList-90 (SCL-90) [30]. As the SCL-90 was not collected at T5 (the 5-year follow-up measurement), this time point will be omitted from analysis. The SCL-90 is a self-report questionnaire for the assessment of psychopathological symptoms in adults, which demonstrated good reliability, validity, and internal consistency [30]. The depression subscale of the SCL-90 contains 16 items with responses measured on a 5-point Likert Scale ranging from 1 (never) to 5 (extremely). Hence, the lowest possible sum score was 16 and the highest was 80. For the trajectory analysis, the total sum score of the SCL-90 depression subscale was used. Based on universal standards for the general population a moderate SCL-90 depression score ranges between 20 and 24 [30]. The depression subscale of the SCL-90 has been validated in various samples [31]–[33].

#### Covariates

At baseline, data on sex, age (younger or older than 21 years), rank, educational level, year of deployment, previous deployments (yes or no), and traumatic experiences in early life were gathered. Rank was categorized into private, corporal, non-commissioned officer, and staff officer. Educational level was divided into three levels: low (some years of high school, but not finished), medium (finished high school), and high (any years of college or university education). Year of deployment was categorized into two groups: 2005-2006 and 2007-2008. Potential early life trauma was assessed with the Early Trauma Inventory Self-Report – Short Form (ETISR-SF), which is a 27-item questionnaire assessing traumatic experiences that have occurred before the age of 18 [34].

At T2, participants reported their function and experiences during the mission. Function described the participants role during the mission as activities inside the base (e.g., logistics), activities outside the base (e.g., patrols), or both. In addition, participants completed the Deployment Experience Scale (DES). The DES is a checklist to assess exposure to 19 combat-related and traumatic stressors during deployment such as witnessing dead, physical injury, and personal danger [5].

At all time points, PTSD symptoms were measured with the Dutch Self-Rating Inventory for PTSD (SRIP), containing 22 questions scored on a 4-point Likert scale ranging from 1 (not at all) to 4 (very much). These items correspond to the DSM-IV criteria for PTSD [35]. The SRIP showed good reliability, validity, and internal consistency [36], [37]. For the indication of substantial PTSD symptoms, a cut-off of 39 was used [38].

**Figure 1.**
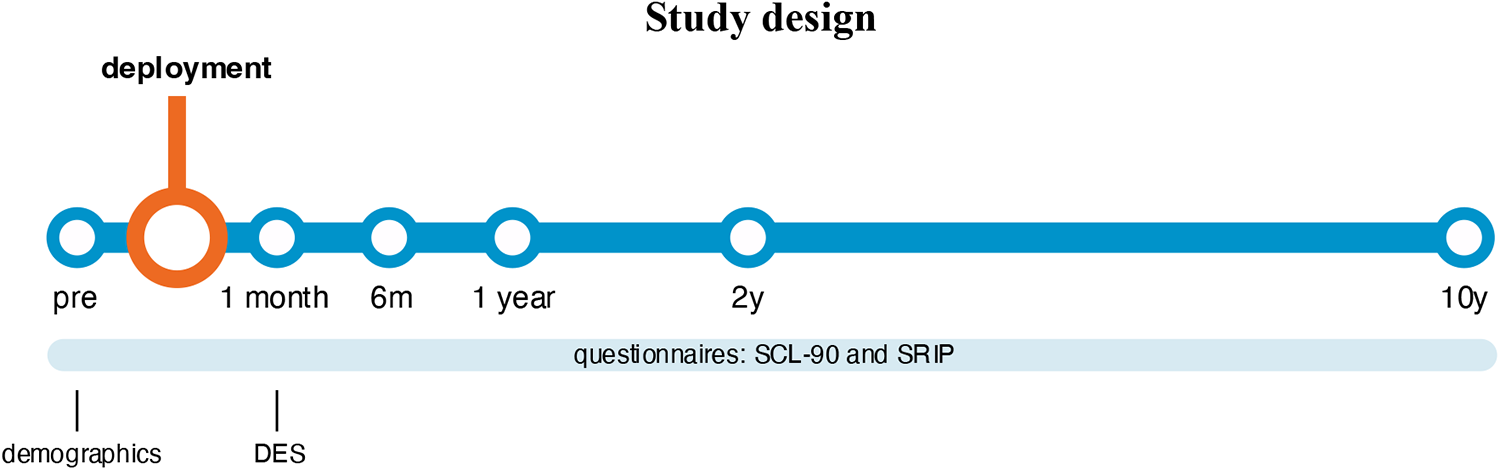
Design of the used part of the Prospective Research in Stress-Related Military Operations (PRISMO). The PRISMO study collected data on 7 time points, but the SCL-90 was not collected at T5 and therefore this time point is omitted. Abbreviations: m, month; y, year; DES, Deployment Experience Scale; SCL-90, Dutch Symptom CheckList-90; SRIP, Self-Rating Inventory for Post-Traumatic Stress Disorder.

### Statistical Analysis

#### Descriptive statistics

Participants with depression data at one or more time point(s) were included in the trajectory analysis. As a control for selection bias, the groups with and without any depression measurement were compared and assessed with chi-square tests. Differences in depression scores over time were assessed with Friedman’s ANOVA. Descriptive statistics are presented as numbers and percentages.

#### Trajectory analysis

The data showed a skewed distribution with inflation at the minimum of the scale. Because preliminary analysis indicated convergence problems of the LCGA model due to this skew, a Box-Cox transformation was performed (Supplementary figure S1). This transformation makes it possible to examine the patterns of heterogeneity between participants in more detail. As LCGA requires complete data, missing data on the depression and PTSD items was imputed using the missForest single imputation algorithm, which uses a random forest model of all other variables to interpolate missing values. In simulation studies, single imputation using missForest shows comparable performance to best practice algorithms [39]. Additionally, it can be applied to all types of data and does not require assumptions about the distribution of the data [39]. As LCGA assumes that data are not multivariate normal, non-parametric imputation is appropriate [40]. Measurement invariance analyses for depressive symptoms are described in Supplementary table S2. For the trajectory analysis and assessment of covariates (demographics, deployment experience and PTSD symptoms) we followed the three-step approach which accounts for classification errors of the model [41]. All analyses were performed using the programming language R (version 4.2.2, package for trajectory analysis: tidySEM; [42]) (with significance level set at *p* < .05 (two-tailed)). All R code is made publicly available on GitHub at https://github.com/XandraPlas/prismo_trajectanalysis

To detect heterogeneity in the development of depressive symptoms after deployment, LCGA was conducted, which assumes the existence of multiple prototypical developmental trajectories [16]–[18]. Previous studies [11] identified three to six subgroups of depression trajectories. For this reason, models with one-to seven classes were estimated. To model the potential effect of deployment on depression, a dummy variable was included that was zero before deployment (T0) and one after deployment (T1-T4, T6). To model potential change in the development of depressive symptoms after deployment, both linear and quadratic terms were included. Differences between the identified trajectories in intercept, slope and step parameters were assessed with Wald tests.

The best performing model was selected based on a combination of the Bayesian information criterion and interpretability [17], [18], [43]. Common fit indices are Information Criteria (IC) including the Bayesian Information Criteria (BIC), sample-size-adjusted BIC (saBIC), and the Akaike Information Criteria (AIC). The general principle is to select the model with the lowest value on these ICs [40], [44], [45]. With contradicting ICs, the inflection point in a scree plot can be used to decide which additional classes have a minor contribution to the decrease in ICs [46]. To assess the model classification performance, entropy, posterior classification probability (probability of belonging to the class one is assigned to), and class size were evaluated. Clearly separable solutions were considered to be desirable because we intended to interpret the class solution. Class separability is related to entropy, with high entropy indicating separate and clearly distinguishable classes [40], [47]. We thus eliminated solutions with entropy < .90 from consideration. Second, if individuals’ posterior classification probability is high for one class and low for the other classes, the classes are distinct, i.e., the model can adequately group individuals with similar trajectories [48]. We accepted solutions with a minimum average posterior classification probability > .90 [47]. Finally, previous research recommended class sizes of at least 50 individuals or 5% of the total sample [47], [49]. We therefore considered classes with at least 50 participants.

#### Exploration of life events

With the same method used for the covariates, we assessed differences in the occurrence of important life events between the different classes. All participants reported their important life events (e.g., divorce, new job, financial problems) at T3-T6.

## Results

### Descriptive statistics

A total of 1032 military men and women participated in the PRISMO cohort. Of this total, 978 were included in this study. Participants were excluded if they were eventually not deployed (n = 25) or did not have a depression measurement at any of the time points (*n* = 29). The comparison between the group with and without any measurements is shown in Table 1 with all baseline characteristics of the participants. No significant differences were found between participants missing all depression measurements and participants with at least one depression measurement, indicating no selection bias when participants without any outcome value were excluded from the analysis. Supplementary table S3 shows the total number of participants per time point and the percentage of participants with low (< 20), average (20 − 23), or high (> 23) levels of depressive symptoms based on the norm of Arrindell & Ettema (2005). The sample average depression score did not significantly change over the 10 years after deployment (χ^2^(5) = 9.56, *p* = .09).

**Table 1.**
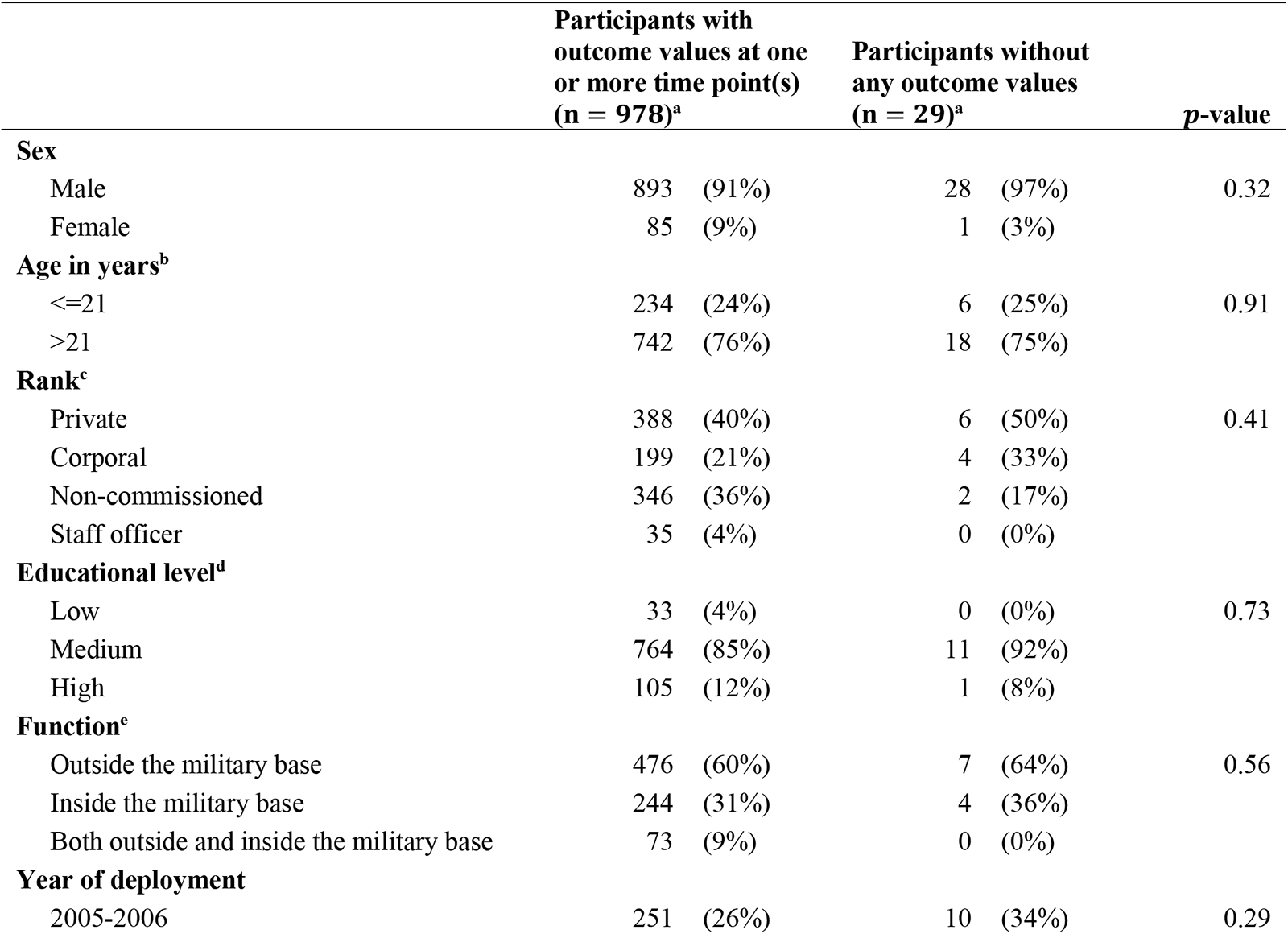
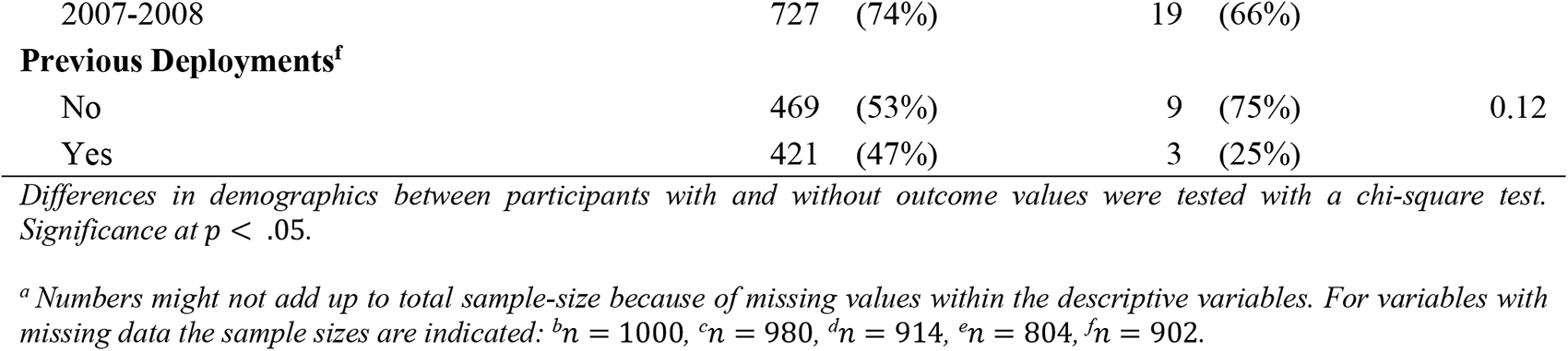
Demographics and baseline characteristics of the deployed participants in the PRISMO cohort, n (%).

### Trajectory analysis

#### Model selection

A complete table of model characteristics and fit statistics of the linear and quadratic models are shown in Supplementary table S4. A selection of characteristics and fit statistics is presented in Table 2 for the linear models. All quadratic models, except the one-class quadratic model, encountered convergence problems and failed to find a final solution.

**Table 2.** Model characteristics and fit indices.

| Model | LL | Parameters | AIC | BIC | saBIC | Entropy | Minimum posterior classification probability | Minimum group size (% of total sample) | LMR | LMR $p$-value |
| --- | --- | --- | --- | --- | --- | --- | --- | --- | --- | --- |
| **Linear step** |  |  |  |  |  |  |  |  |  |  |
| 1 | 526 | 9 | -1035 | -991 | -1049 | **1.00** | **1.00** | **100%** |  |  |
| 2 | 1903 | 13 | -3779 | -3716 | -3802 | **0.94** | **0.97** | **24%** | 2626 | **<0.01** |
| 3 | 2251 | 17 | -4468 | -4385 | -4498 | **0.93** | **0.93** | **9%** | 665 | **<0.01** |
| 4 | 2351 | 21 | -4660 | -4558 | -4699 | **0.94** | **0.90** | **6%** | 191 | **<0.01** |
| 5 | 2431 | 25 | -4811 | -4689 | -4858 | **0.90** | 0.83 | **6%** | 152 | **<0.01** |
| 6 | 2466 | 29 | -4874 | -4732 | -4928 | **0.90** | 0.83 | 2% | 68 | **<0.01** |
| 7 | 2484 | 33 | -4903 | -4742 | -4965 | **0.91** | 0.80 | 1% | 35 | **<0.01** |

*Bold is in accordance with our criteria or indicates significance ( $p < .05$ ).*
*Abbreviations: LL, Log-likelihood; AIC, Akaike Information Criteria; BIC, Bayesian Information Criteria; saBIC, sample-size-adjusted Bayesian Information Criteria; LMR, Lo-Mendell-Rubin likelihood test.*

For the linear models, a reduction in BIC, saBIC and AIC was observed with increasing class number (Supplementary figure S5). The reduction in ICs flattened after three classes. In addition, the Lo-Mendell-Rubin (LMR) likelihood ratio test was significant with every additional class and entropy was above . 90 for all models. Posterior group probability was acceptable (above . 90) up to the four-class model. Therefore, the models with five or more classes were not accepted. For all models with one to four classes, the class size was larger than 5% and consisted of at least 50 individuals.

Altogether, most quadratic models did not converge, and both the three-and four-class model performed well and had acceptable classification performance. But the four-class model was able to capture the small group of soldiers exhibiting a late-onset increase in depressive symptoms. Therefore, we selected the linear four-class model as the final model.

#### Depression trajectories

The four classes identified were a resilient class with 639 participants (65%), an intermediate-stable class with 199 participants (20%), a symptomatic-chronic class with 86 participants (9%), and a late-onset-increasing class with 54 participants (6%) (Figure 2). The resilient group showed low levels of depressive symptoms before deployment with a small significant decrease after deployment, followed by stability (*p* < .01, Supplementary table S6). The intermediate-stable and symptomatic-chronic group showed a significant increase in depressive symptoms after deployment that slightly decreased over time for the intermediate-stable group and remained elevated and stable until the 10-year time point for the symptomatic-chronic group (*p* < .01, Supplementary table S6). The late-onset-increasing group started between the low and intermediate group before deployment, showed a significant decrease in depressive symptoms from pre-to post-deployment followed by an increase from there until 10 years post-deployment (*p* < .01, Supplementary table S6).

Results of the Wald tests (Supplementary table S7) showed that the pre-deployment scores for depressive symptoms (intercepts) and the increase from pre-to 1-month post-deployment (step parameter) differed significantly between all classes (*p* < .01). Furthermore, the trajectories differed significantly in slope (*p* < .01). (See Supplementary table S8 for pairwise comparisons with Bonferroni correction).

**Figure 2.**
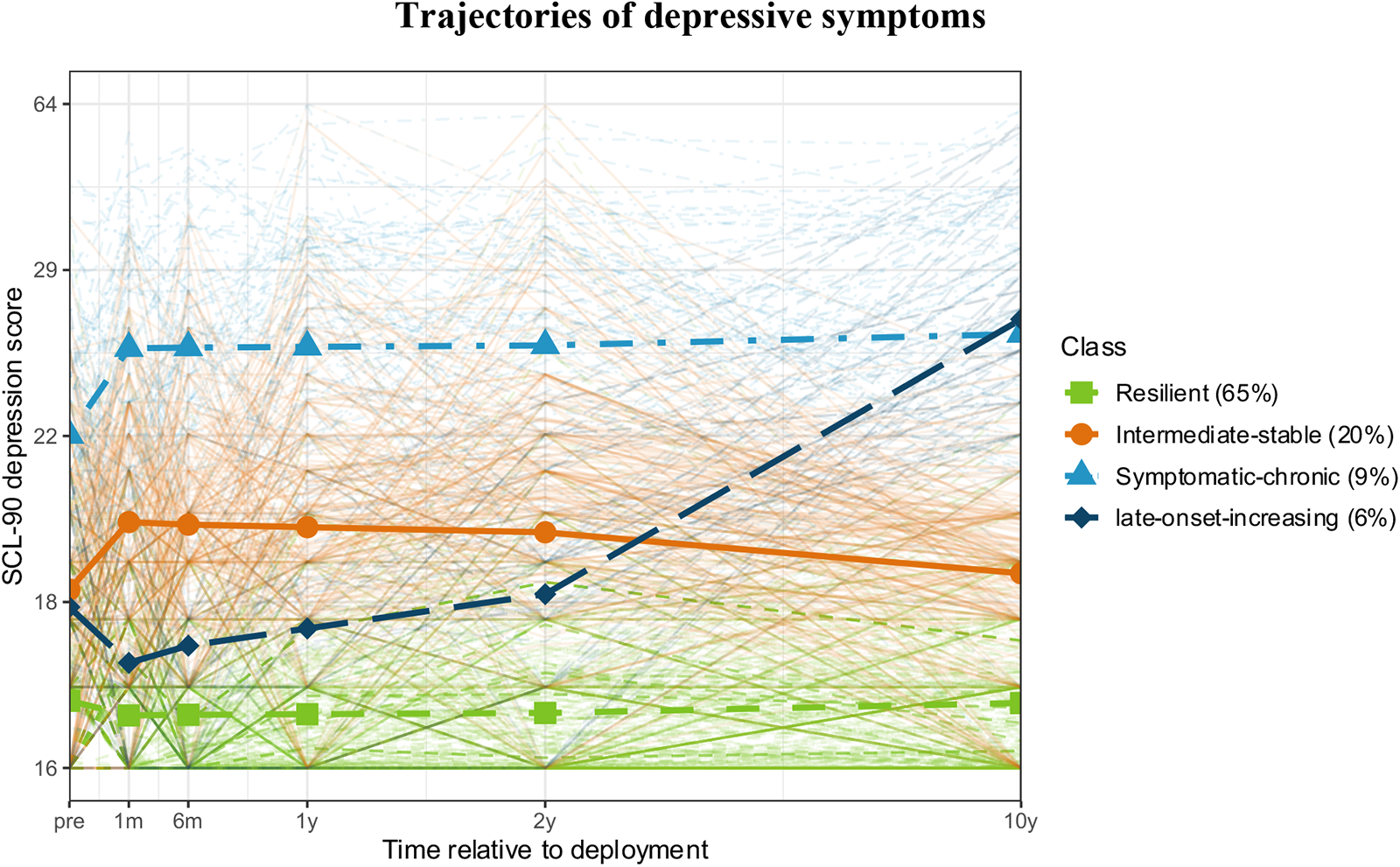
Predicted depression trajectories and individual depression scores at the different time points. Different colors indicate the four different classes. SCL-90 (Short CheckList – 90) score is rescaled from Box-Cox transformation.

### Covariates

Assessment of covariates using the three-step approach revealed that the late-onset-increasing group had a higher proportion of soldiers under 21 years compared to the resilient, intermediate-stable, and symptomatic-chronic group (respectively, Δ*LL*(1) = 12.5, Δ*LL*(1) = 5.1, Δ*LL*(1) = 9.7, *p* < .05). The trajectories did not differ in sex (Δ*LL*(5) = 6.3, *p* = .27), rank (Δ*LL*(11) = 14.0, *p* = .26), educational level (Δ*LL*(9) = 7.1, *p* = .63), function (Δ*LL*(8) = 8.1, *p* = .43), year of deployment (Δ*LL*(5) = 3.6, *p* = .61), or previous deployments (Δ*LL*(6) = 3.0, *p* = .81). For early life traumas, the symptomatic-chronic group experienced more early life traumas compared to the other trajectories (*p* < .01) and both the resilient and late-onset-increasing group experienced significantly less early life traumas compared to the intermediate-stable group (ΔLL(2) = 21.6, p < .01; ΔLL(2) = 8.1, p < .05) (Supplementary figure S9).

Significant disparities in deployment stressors were observed in five stressors of the DES, with the resilient group exhibiting lower percentages of deployment stressors compared to other groups (Table 3). Soldiers in the resilient group reported a lower frequency of personal danger and fewer instances of feeling a sense of mission uselessness compared to soldiers in the intermediate-stable group (Δ*LL*(1) = 4.3, *p* < .05; Δ*LL*(1) = 13.1, *p* < .01) and the symptomatic-chronic group (Δ*LL*(1) = 13.9, Δ*LL*(1) = 8.6, *p* < .01). Additionally, they reported ‘hearing injured scream’ less frequently than the intermediate-stable group (Δ*LL*(1) = 7.8, *p* = < .01). Finally, the resilient group was less likely to encounter insufficient opportunities to intervene and reported fewer instances of feeling out of control compared to the intermediate-stable group (Δ*LL*(1) = 10.1, Δ*LL*(1) = 27.4 *p* < .01;), the symptomatic-chronic group (Δ*LL*(1) = 16.9, Δ*LL*(1) = 21.9, *p* < .01), and the late-onset-increasing group (Δ*LL*(1) = 10.8, *p* < .01; Δ*LL*(1) = 6.3, *p* < .05).

**Table 3.** Five deployment stressors from the DES that significantly differed between the trajectories.

|  | **Resilient**<br>**(65%)** | **Intermediate-**<br>**stable (20%)** | **Symptomatic**<br>**-chronic (9%)** | **Late-**<br>**onset-**<br>**increasing**<br>**(6%)** |
| --- | --- | --- | --- | --- |
| Personal danger | 25%<sup>ab</sup> | 34%<sup>a</sup> | 47%<sup>b</sup> | 35% |
| Screaming of injured | 17%<sup>a</sup> | 27%<sup>a</sup> | 24% | 27% |
| Insufficient opportunities to intervene | 12%<sup>abc</sup> | 23%<sup>a</sup> | 32%<sup>b</sup> | 30%<sup>c</sup> |
| Insufficient control over situation | 8%<sup>abc</sup> | 23%<sup>a</sup> | 28%<sup>b</sup> | 20%<sup>c</sup> |
| Mission experienced as useless | 10%<sup>ab</sup> | 21%<sup>a</sup> | 23%<sup>b</sup> | 18% |

*Same superscripts indicate significant different percentages within that row ( $p < .05$ ). For example, within the resilient group, 25% reported personal danger. This was significantly lower than the intermediate-stable group (both an ‘a’ as superscript) and the symptomatic-chronic group (both a ‘b’ as superscript). The intermediate-stable and symptomatic-chronic group did not differ significantly (they do not share the same superscript) and the late-onset-increasing group was not significantly different from any other group (no superscript).*

#### Depression and PTSD

Significant differences in PTSD symptom scores were observed across all four trajectory groups at nearly every time point (Figure 3, Supplementary table S10). The resilient group consistently exhibited significantly lower levels of PTSD symptoms compared to the intermediate-stable, symptomatic-chronic, and late-onset-increasing group (*p* < .01). Additionally, the symptomatic-chronic group scored higher on PTSD symptoms compared to the intermediate-stable and late-onset-increasing group across all time points (*p* < .01). Finally, the late-onset-increasing group reported significant different levels of PTSD symptoms compared to the intermediate-stable group (*p* < .01) at all time points, except for the 1-year time point (*p* = .06).

**Figure 3.**
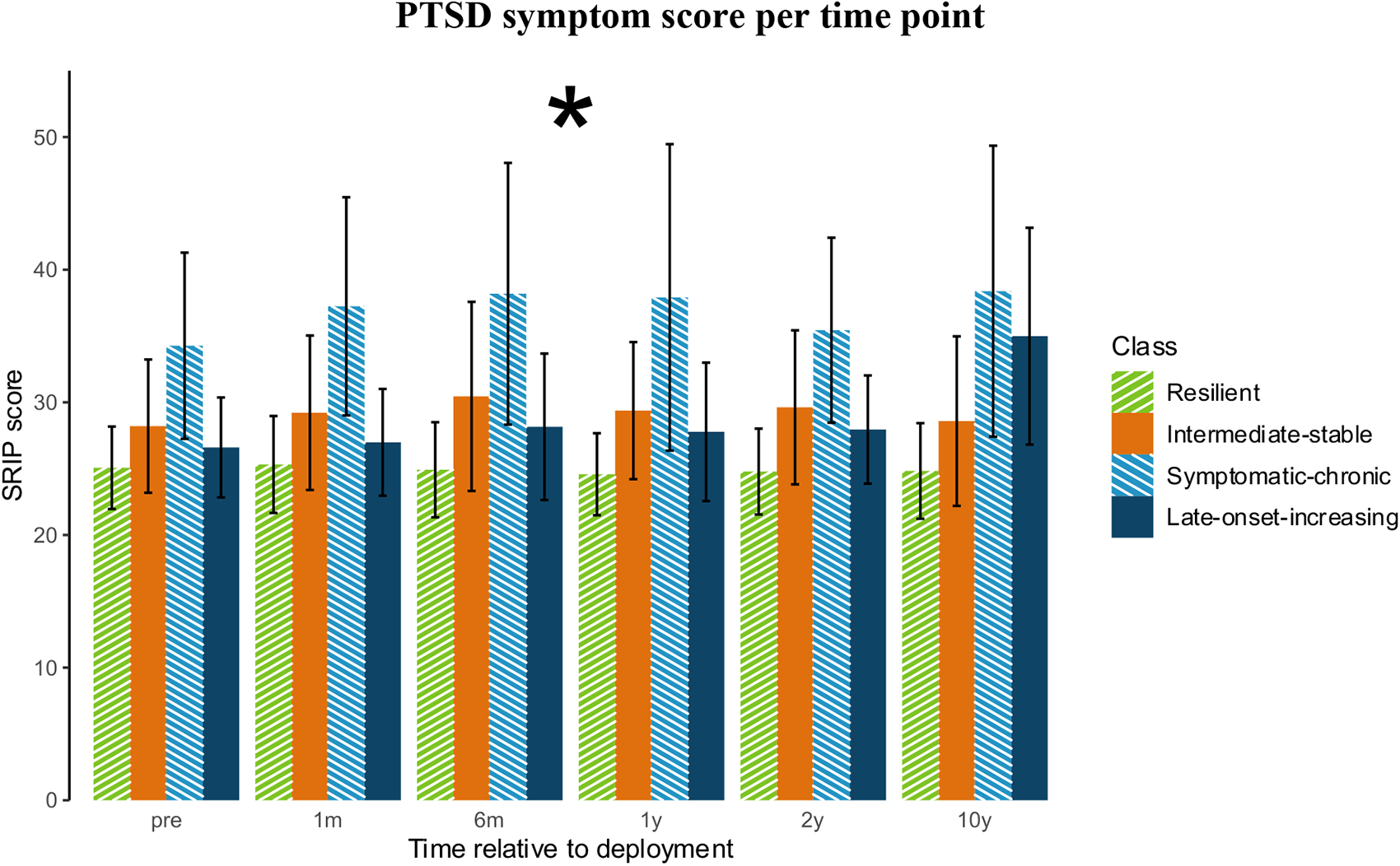
PTSD scores per time point (mean and standard deviation (n = 978)). Colors indicate different classes. *All trajectories differed significantly at each time point (*p* < .05), except the intermediate-stable and late-onset-increasing group at the 1-year time point (*p* = .064).

#### Depression and important life events

From deployment to the 1-year time point, only participants in the resilient group reported fewer life events compared to the symptomatic chronic group (*p* < .05) (Supplementary figure S11). Starting from two years after deployment and continuing to the 10-year time point, the resilient group consistently exhibited fewer life events compared to all the other trajectories (*p* < .05) (Supplementary figure S11). Trajectories differed for example in being sick, relationship problems or sexual problems (Supplementary table S12-S14).

## Discussion

The current study investigated trajectories in the development of depressive symptoms up to 10 years after deployment in military personnel. Therefore, a LCGA was performed based on a group of 978 veterans who were deployed as part of ISAF. In line with the hypothesis, four distinct trajectories were identified: 1) resilient, 2) intermediate-stable, 3) symptomatic-chronic, and 4) late-onset-increasing. The trajectories differed in the proportion of individuals younger than 21 years, deployment stressors, and PTSD symptom scores.

Most of the study sample fell within the resilient trajectory (65%), displaying no or low levels of depressive symptoms. This finding is consistent with previous studies conducted in military samples, where a resilient group (55.9% – 86.5%) was observed comprising most of the participants [12]–[14]. Furthermore, this large resilient group was also identified in recent studies investigating depression trajectories in students, older adults, and adolescents[7]–[9]. The intermediate-stable trajectory (20%) exhibited an increase in depressive symptoms after deployment and slightly elevated levels of depressive symptoms compared to the resilient trajectory. This intermediate stable trajectory with low levels of depression was also found in previous studies in the civilian population [50], [51]. Both the resilient and intermediate-stable trajectories remained below average symptom levels (average levels: 20 − 23 [30]. These findings support the idea that resilience is more common than expected [52] and might imply resiliency in deployed military personnel, which is a protective factor for depression [53], [54]. In addition, assuming that deployed military personnel are an extraordinary selection of young and healthy individuals, high resilience in the face of adversity is expected.

Unlike the resilient and intermediate-stable trajectories, a symptomatic-chronic group (9%) was present which showed medium levels of symptoms before deployment, and this increased to high levels (> 23 [30]) after deployment. This finding is in line with the studies of Sampson and colleagues [13], [14], who found a chronic (dysfunction) group within a military cohort, consisting of respectively 12.6% and 9.2% of the individuals. Although trajectories with high depressive symptoms over time are rare, studies both in the military and civilian samples report long-term chronic, high levels of depressive symptoms in small proportions (<10%) of the study population [11], [13], [14].

Finally, the model was able to capture a late-onset-increasing group (6%). This increasing group has also been identified in previous military studies [12], [13], [55]. Our trajectory resembled the low-increasing group discovered by Karstoft and colleagues [12], although there was a notable distinction in the timing of the increase. In their study, the trajectory drastically increased directly at homecoming, whereas ours increased more gradually. On the other hand, Sampson and colleagues found a mild increasing group, which differed from our late-onset-increasing group that experienced a large increase of symptoms. But looking at our results, this disparity can be explained by the shorter study period of Sampson and colleagues. It was only towards the 10-year point that our late-onset-increasing group exhibited above-average levels of depressive symptoms.

Significant differences between all trajectories were found for both the intercept, step (from pre-to 1-month post), and slope, indicating that individuals at risk might be identified before deployment and react differently to deployment. It may be speculated that the differences in the trajectories of depressive symptoms are influenced by multiple factors, including pre-existing individual differences and the experience of significant life events. Our results showed that individuals in trajectories with higher depression scores, were more likely to experience important life events. It is important to note, however, that these results cannot be interpreted causally because life events were measured during the course of the study. It is thus equally plausible that these life events influenced trajectories of depressive symptoms, or that trajectories of depressive symptoms influenced the likelihood of experiencing, or perceived salience, of life events. Furthermore, the significant differences in intercepts between classes suggest that individuals may have varying levels of vulnerability or predispositions to depression even before deployment. These vulnerabilities could be attributed to genetic factors, early-life experiences, personality traits, or other underlying biological and psychological factors [55]–[57]. If individuals with higher vulnerability or pre-existing depressive symptoms enter the military, the exposure to stressful and traumatic events during deployment (e.g., separation from loved ones, combat, or witnessing injuries and harmful situations) could exacerbate depressive symptoms, leading to a more severe trajectory of depressive symptoms post-deployment. Further research is needed to understand the interplay between pre-existing vulnerabilities, exposure to deployment stressors, and the development of depressive symptoms.

Analysis of covariates revealed no differences between trajectories for demographic variables, except for age proportions. While others have found female veterans to be more likely to develop depression [58], we did not find any differences in sex between the trajectories. This could be explained by lack of power, as our sample included only a small number of women. The trajectories did differ in the number of traumatic events in early life, with the symptomatic-chronic group reporting the highest number. This aligns with the study by Sampson and colleagues [14], and a previous PRISMO study have also found that early trauma is negatively associated with aspects of personality, which in turn leads to a higher risk of adult psychopathology[59]

Regarding deployment stressors and PTSD, differences were observed among distinct trajectories. Previous research has established a consistent correlation between depression and military deployment, particularly in deployments that involve combat exposure. Conversely, military deployment without such exposure has been linked to reduced risk of depression [3], [4], [60]. In our study, those who exhibited a resilient trajectory tended to report fewer negative experiences during deployment, such as personal danger, screaming of injured, and feeling out of control. Again, it is important to note that these results cannot be interpreted causally. Considering the differences of depressive symptoms between the trajectories prior to deployment, it is plausible to propose that these trajectories of depressive symptoms influenced the perception of stressors during deployment. In accordance with our results, Porter and colleagues [61] found that depression was associated with witnessing abuse, feeling in danger, knowing someone is injured, or being injured oneself. Regarding PTSD, trajectories with higher depressive symptoms also exhibited higher levels of PTSD symptom. Notably, the symptomatic-chronic and late-onset-increasing group approached the PTSD cut-off score of 39 [38] at the 6-month, 1-year, and 10-year follow-up assessments. However, a trend emerged indicating that participants with higher depressive symptoms had higher PTSD symptom scores. PTSD and depression co-occur frequently [24], and while there are overlapping symptoms, PTSD and depression are distinguishable disorders characterized by unique patterns of symptoms [62], [63].

One of the limitations of the current study is the use of self-report questionaries, which might not correspond with clinician assessment of depression. However, for measures of depression, there remains a lack of consensus regarding the extent of disagreement between clinician-rated and self-reported measures of depression [64]–[66]. Thereby, the use of a self-report questionnaire was consistent over all time points, ensuring that symptoms are consistently assessed in the same way. This was also confirmed with the analysis of measurement invariance. In addition, there was a lack of diversity in the sample population, with a majority of participants being of the same ethnicity and a minority of women. This limits the generalizability of the study’s findings to other groups. Another limitation is that around 40% of participants did not complete all measurements. This is a common limitation in longitudinal studies, and we cannot discount the possibility that nonresponse may have an influence on the study results. Additionally, most criteria for model selection in LCGA have known limitations [40], [47], resulting in subjective judgements and inconsistencies among different individuals or groups. We have chosen strict criteria, ensuring that the performance of the model was appropriate. At last, the absence of information regarding timing and type of received treatment in the study period limits interpretability. As a result, we cannot attribute any changes in trajectories to the treatments they may have received during the study period. One strength of this study is that it was one of the first to perform LCGA using free open-source software, most notably R and the tidySEM package. Furthermore, all code used in this analysis is available in a reproducible repository, thereby enabling all researchers to reuse the code to perform similar analyses.

## Conclusion

In conclusion, four trajectories were identified in the development of depressive symptoms in a military sample up to 10 years after deployment. The late-onset-increasing group had a higher proportion of individuals under 21 years compared to the resilient and symptomatic group. Additionally, the resilient group was less likely to experience deployment stressors. Between all trajectories PTSD symptom scores differed significantly at nearly every time point, with the symptomatic-chronic trajectory exhibiting the highest PTSD symptom scores.

The findings of this study are consistent with previous research on depression trajectories in various populations. The majority fell within the resilient trajectory, indicating that resilience is more common than expected and supporting the notion that deployed military personnel possess a high level of resilience as a protective factor against depression. These insights in the long-term development of depressive symptoms may guide decisions regarding deployment, mental health care, and policies within the army. However, the question remains why depressive symptoms develop differently across military personnel and veterans. More research is needed to investigate which factors are associated with and predict the different depression trajectories.

## Supporting information

Supplementary material_Plas et al

## Data availability

The data that support the findings of this study are available upon reasonable request from any qualified investigator. Restrictions apply to the availability of these data due to privacy or ethical considerations. For inquiries regarding data access, please contact the Brain Research and Innovation Centre at.

## Conflicts of interest

All authors declare no potential conflicts of interest with respect to the research, authorship, and/or publication of this article.

## Funding statement

This work was supported by the Dutch Ministry of Defence. The funder had no role in the design and reporting of the study.

## Acknowledgments

The authors thank the Dutch commanders and troops and all members of the PRISMO team involved in data acquisition for their ongoing commitment to the study.

## Notes

### Competing Interest Statement

The authors have declared no competing interest.

### Author Declarations

Institutional Review Board of the University Medical Centre Utrecht (Utrecht, The Netherlands) gave ethical approval of this work.

### Summary of Updates

In this revised file the incorrect in-text references to supplementary figures/tables are corrected and a typo in the email address for a data request has been corrected.

