## Supplementary material_Plas et al for "Long-term Trajectories of Depressive Symptoms in Deployed Military Personnel: A 10-year prospective study"

*Transformations*

Multiple transformations were compared to reduce the skew: square (sqr), cube root (sqrt), log, and Box-Cox (Supplementary figure S1). The Box-Cox transformation was able to best reduce the skewness.

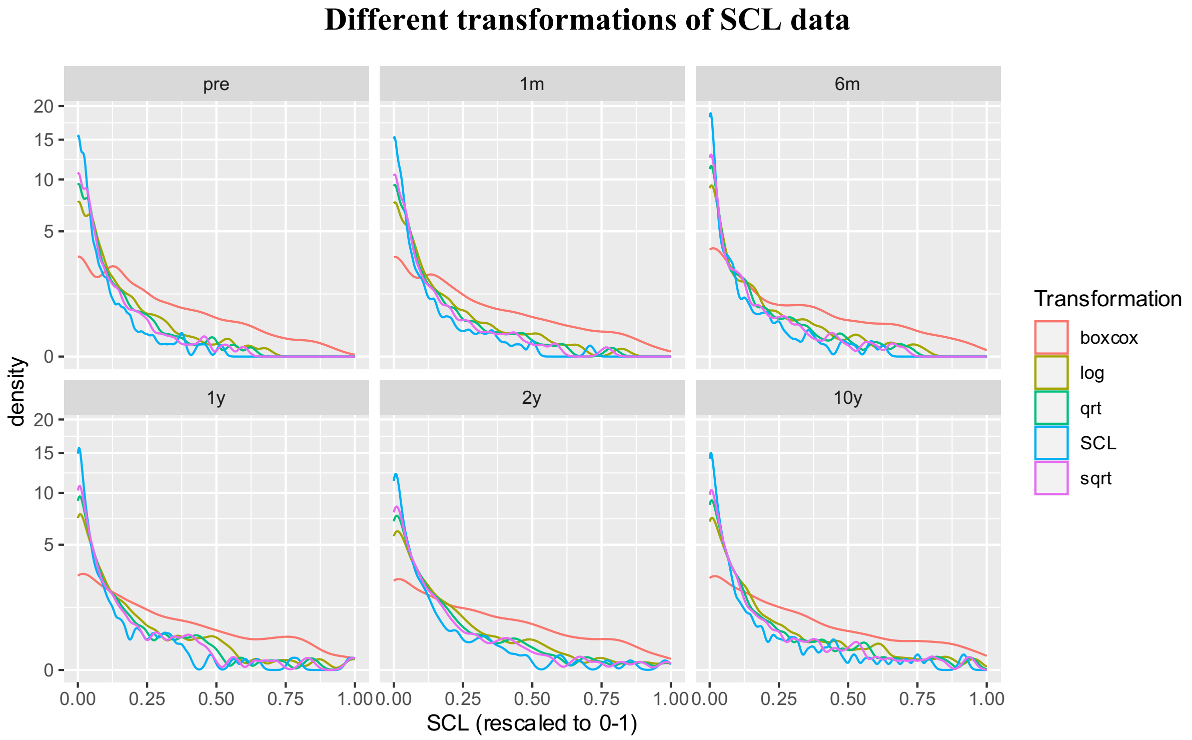


***Supplementary figure S1*** *– Multiple transformations on SCL-90 depression data per measurement round. Different colors indicate different methods for data transformation.*

*Measurement invariance of SCL-90-R depression data*

Measurement invariance was assessed with Confirmatory Factor Analysis (CFA). The results are presented in Supplementary table S2. In general, larger Root Mean Square Error of Approximation (RMSEA) and smaller Comparative Fit Index (CFI) or Tucker-Lewis Index (TLI) indicate worse model fit, but the literature suggests cut-off criteria [66]. The criterium for CFI and TLI was > .95 and for RMSEA < .05.

**Supplementary table S2**

Fit indices for measurement invariance assessment.

| Name | Parameters | Chi-square | df | CFI | TLI | RMSEA | SRMR |
|---|---|---|---|---|---|---|---|
| **CFA** | 79 | 495 | 104 | **1.00** | **1.00** | **0.03** | 0.04 |
| **Configural** | 434 | 7662 | 4449 | **0.99** | **0.99** | **0.03** | 0.10 |
| **Metric** | 354 | 11673 | 4529 | **0.98** | **0.98** | **0.04** | 0.12 |

*Bold is in accordance with our criteria.*

*Abbreviations: CFA, Confirmatory Factor Analysis; df, degrees of freedom; CFI, Comparative Fit Index; TLI, Tucker-Lewis index; RMSEA, Root Mean Square Error of Approximation; SRMR, Standardized Root Mean-square Residual.*

### *Scores on depressive symptoms*

**Supplementary table S3**

Number of participants with low, medium, or high levels of depressive symptoms, n (%).

| | Depression low | | Depression medium | | Depression high | | n total |
|---|---|---|---|---|---|---|---|
| Pre-deployment | 678 | (81%) | 122 | (15%) | 34 | (4%) | 834 |
| 1 month | 646 | (79%) | 112 | (14%) | 55 | (7%) | 813 |
| 6 months | 579 | (79%) | 96 | (13%) | 60 | (8%) | 735 |
| 1 year | 421 | (75%) | 77 | (14%) | 61 | (11%) | 559 |
| 2 years | 394 | (72%) | 96 | (18%) | 58 | (11%) | 548 |
| 10 years | 455 | (76%) | 80 | (13%) | 67 | (11%) | 608 |

*Depression was assessed with the SCL-90. Cut-off <20 for symptom levels below average, 20-23 for average symptom levels and >23 for levels above average* [29].

*Model characteristics and fit statistics*

**Supplementary table S4**
Model characteristics and fit statistics of the linear and quadratic models.

| Model | LL | Parameters | AIC | BIC | saBIC | Entropy | Minimum posterior classification probability | Maximum posterior classification probability | Minimum group size (% of total sample) | Maximum group size (% of total sample) | LMR | LMR *p*-value | Convergence problem |
|---|---|---|---|---|---|---|---|---|---|---|---|---|---|
| **Linear** | | | | | | | | | | | | | |
| 1 | 526 | 9 | -1035 | -991 | -1049 | **1.00** | **1.00** | 1.00 | **100%** | 100% | | | |
| 2 | 1903 | 13 | -3779 | -3716 | -3802 | **0.94** | **0.97** | 0.99 | **24%** | 76% | 2626 | **<0.01** | |
| 3 | 2251 | 17 | -4468 | -4385 | -4498 | **0.93** | **0.93** | 0.98 | **9%** | 67% | 665 | **<0.01** | |
| 4 | 2351 | 21 | -4660 | -4558 | -4699 | **0.94** | **0.90** | 0.99 | **6%** | 65% | 191 | **<0.01** | |
| 5 | 2431 | 25 | -4811 | -4689 | -4858 | **0.90** | 0.83 | 0.98 | **6%** | 61% | 152 | **<0.01** | |
| 6 | 2466 | 29 | -4874 | -4732 | -4928 | **0.90** | 0.83 | 0.98 | 2% | 60% | 68 | **<0.01** | |
| 7 | 2484 | 33 | -4903 | -4742 | -4965 | **0.91** | 0.80 | 0.98 | 1% | 60% | 35 | **<0.01** | |
| **Quadratic** | | | | | | | | | | | | | |
| 1 | 536 | 10 | -1052 | -1003 | -1068 | **1.00** | **1.00** | 1.00 | **100%** | 100% | | | |
| 2 | 1932 | 15 | -3834 | -3761 | -3861 | **0.94** | **0.97** | 0.99 | **24%** | 76% | 2663 | **<0.01** | TRUE |
| 3 | 2295 | 20 | -4550 | -4452 | -4586 | **0.93** | **0.94** | 0.98 | **9%** | 66% | 692 | **<0.01** | TRUE |
| 4 | 2387 | 25 | -4723 | -4601 | -4769 | 0.88 | 0.84 | 0.97 | **6%** | 61% | 175 | **<0.01** | TRUE |
| 5 | 2479 | 30 | -4898 | -4752 | -4955 | **0.90** | 0.82 | 0.98 | **6%** | 61% | 177 | **<0.01** | TRUE |
| 6 | 2534 | 35 | -4998 | -4827 | -5064 | 0.89 | 0.77 | 0.97 | 3% | 58% | 105 | **<0.01** | TRUE |
| 7 | 2575 | 40 | -5070 | -4875 | -5146 | 0.89 | 0.80 | 0.95 | 2% | 55% | 78 | **<0.01** | TRUE |

*Bold is in accordance with our criteria or indicates significance ($p < .05$). If the model encountered convergence problems, the best fit solution is presented.*
*Abbreviations: LL, Log-likelihood; AIC, Akaike Information Criteria; BIC, Bayesian Information Criteria; saBIC, sample-size-adjusted Bayesian Information Criteria; LMR, Lo-Mendell-Rubin likelihood test.*

*Information Criteria – AIC, BIC, saBIC*

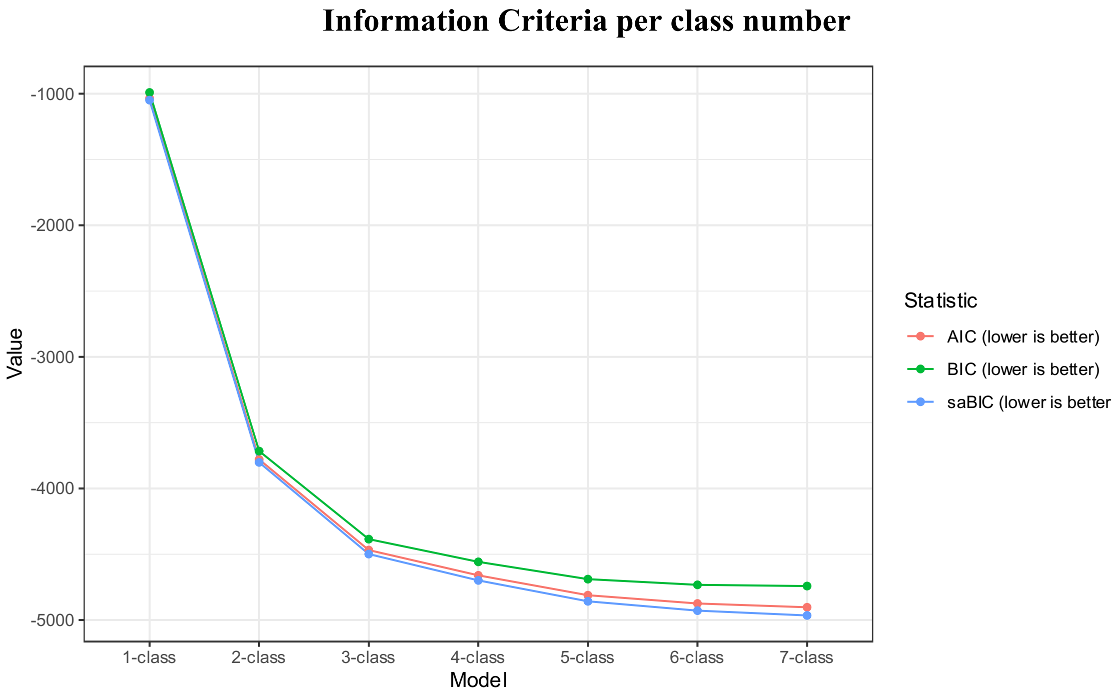


***Supplementary figure S5 -*** *Scree plot with fit statistics of linear models. Colors indicate different fit statistics: AIC, Akaike Information Criteria; BIC, Bayesian Information Criteria; saBIC, sample-size-adjusted BIC.*

### *Model parameters*

**Supplementary table S6**

Model parameters of the final four-class linear model.

| label | Estimated significance | Standard error | *p*-value | Confidence interval | Class |
|---|---|---|---|---|---|
| mix4.weights[1,2] | 0.32*** | 0.03 | **<0.01** | [0.26, 0.37] | |
| mix4.weights[1,3] | 0.13*** | 0.02 | **<0.01** | [0.10, 0.17] | |
| mix4.weights[1,4] | 0.09*** | 0.01 | **<0.01** | [0.06, 0.11] | *NA* |
| Variances.scl1 | 0.02*** | 0.00 | **<0.01** | [0.02, 0.02] | Class 1 |
| Variances.scl2 | 0.02*** | 0.00 | **<0.01** | [0.02, 0.02] | Class 1 |
| Variances.scl3 | 0.01*** | 0.00 | **<0.01** | [0.01, 0.02] | Class 1 |
| Variances.scl4 | 0.02*** | 0.00 | **<0.01** | [0.02, 0.02] | Class 1 |
| Variances.scl5 | 0.03*** | 0.00 | **<0.01** | [0.02, 0.03] | Class 1 |
| Variances.scl6 | 0.02*** | 0.00 | **<0.01** | [0.02, 0.02] | Class 1 |
| Means.i | 0.10*** | 0.01 | **<0.01** | [0.09, 0.11] | Class 1 |
| Means.step | -0.02*** | 0.01 | **<0.01** | [-0.04, -0.01] | Class 1 |
| Means.s | 0.00** | 0.00 | **0.01** | [0.00, 0.00] | Class 1 |
| Means.i | 0.27*** | 0.01 | **<0.01** | [0.25, 0.29] | Class 2 |
| Means.step | 0.10*** | 0.01 | **<0.01** | [0.08, 0.13] | Class 2 |
| Means.s | -0.00*** | 0.00 | **<0.01** | [-0.01, -0.00] | Class 2 |
| Means.i | 0.50*** | 0.02 | **<0.01** | [0.47, 0.53] | Class 3 |
| Means.step | 0.13*** | 0.02 | **<0.01** | [0.09, 0.17] | Class 3 |
| Means.s | 0.00 | 0.00 | 0.29 | [-0.00, 0.00] | Class 3 |
| Means.i | 0.24*** | 0.02 | **<0.01** | [0.20, 0.29] | Class 4 |
| Means.step | -0.08*** | 0.03 | **<0.01** | [-0.13, -0.03] | Class 4 |
| Means.s | 0.03*** | 0.00 | **<0.01** | [0.02, 0.03] | Class 4 |

*Bold indicates significance ($p < .05$).*

**Supplementary table S7**

Results of Wald tests.

| | **df** | **Chi-square** | ***p*-value** |
|---|---|---|---|
| Mean intercept | 3 | 615 | **<0.01** |
| Mean step | 3 | 129 | **<0.01** |
| Mean slope | 3 | 450 | **<0.01** |

*Bold indicates significance (p < .05).*

*Abbreviations: df, degrees of freedom.*

**Supplementary table S8**

Results of pairwise comparison with Wald test of model parameters between classes.

| **Comparison** | | **df** | **Chi-square** | ***p*-value** |
|---|---|---|---|---|
| **Intercept** | | | | |
| Resilient | Intermediate-stable | 1 | 157.0 | **<0.01** |
| | Symptomatic-chronic | 1 | 517.9 | **<0.01** |
| | Late-onset-increasing | 1 | 33.7 | **<0.01** |
| Intermediate-stable | Symptomatic-chronic | 1 | 131.3 | **<0.01** |
| | Late-onset-increasing | 1 | 0.9 | 1.00 |
| Symptomatic-chronic | Late-onset-increasing | 1 | 80.0 | **<0.01** |
| **Step** | | | | |
| Resilient | Intermediate-stable | 1 | 73.0 | **<0.01** |
| | Symptomatic-chronic | 1 | 59.7 | **<0.01** |
| | Late-onset-increasing | 1 | 5.6 | 0.53 |
| Intermediate-stable | Symptomatic-chronic | 1 | 1.6 | 0.61 |
| | Late-onset-increasing | 1 | 41.1 | **<0.01** |
| Symptomatic-chronic | Late-onset-increasing | 1 | 46.0 | **<0.01** |
| **Slope** | | | | |
| Resilient | Intermediate-stable | 1 | 41.0 | **<0.01** |
| | Symptomatic-chronic | 1 | 0.0 | 1.00 |
| | Late-onset-increasing | 1 | 311.8 | **<0.01** |
| Intermediate-stable | Symptomatic-chronic | 1 | 14.6 | **<0.01** |
| | Late-onset-increasing | 1 | 449.1 | **<0.01** |
| Symptomatic-chronic | Late-onset-increasing | 1 | 201.0 | **<0.01** |

*Bold indicates significance (p < .05). p-values are corrected for multiple comparison with Bonferroni correction.*

*Abbreviations: df, degrees of freedom.*

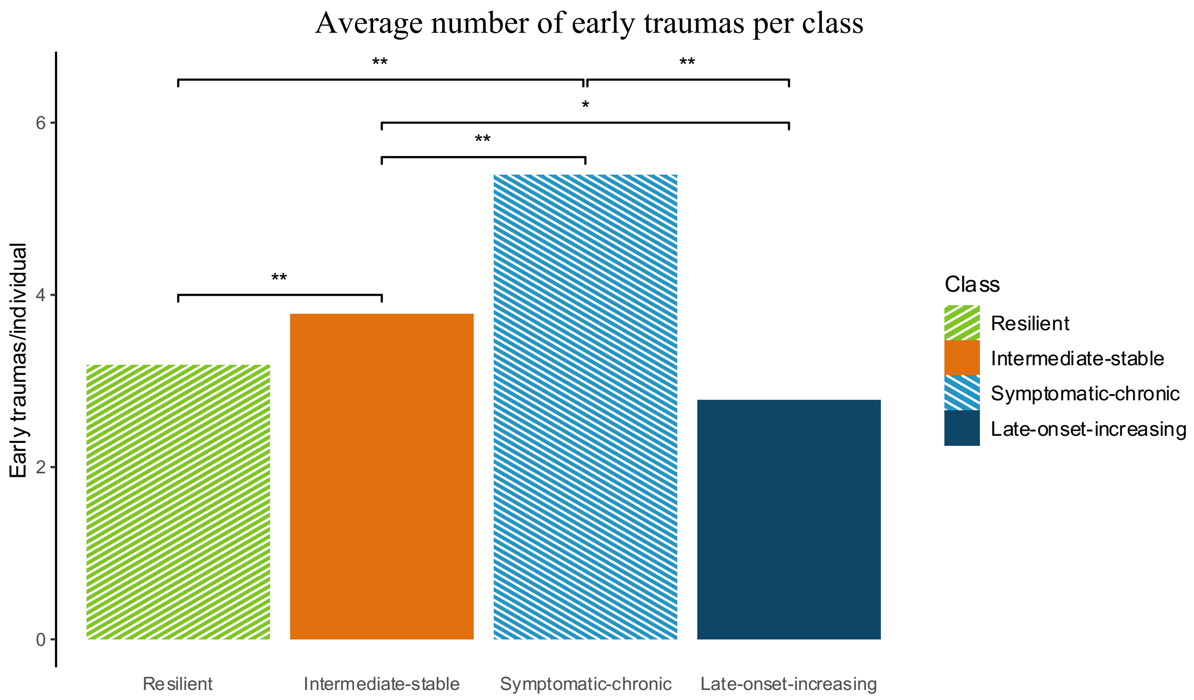


***Supplementary figure S9*** *– Figure to illustrate the average number of early traumas per class. Significant differences are presented with an asterisk* ($* \, p < .05$, $** \, p < .01$).

### *Depression and PTSD*

**Supplementary table S10**

Results of auxiliary models to assess the association between PTSD and depressive symptoms.

| **Comparison** | | **df** | **Δ*LL*** | ***p*-value** |
|---|---|---|---|---|
| **Pre** | | | | |
| Resilient | Intermediate-stable | 2 | 200.8 | **<0.01** |
| | Symptomatic-chronic | 2 | 524.8 | **<0.01** |
| | Late-onset-increasing | 2 | 14.6 | **<0.01** |
| Intermediate-stable | Symptomatic-chronic | 2 | 80.9 | **<0.01** |
| | Late-onset-increasing | 2 | 14.1 | **<0.01** |
| Symptomatic-chronic | Late-onset-increasing | 2 | 77.1 | **<0.01** |
| **1-month** | | | | |
| Resilient | Intermediate-stable | 2 | 207.7 | **<0.01** |
| | Symptomatic-chronic | 2 | 583.2 | **<0.01** |
| | Late-onset-increasing | 2 | 10.1 | **<0.01** |
| Intermediate-stable | Symptomatic-chronic | 2 | 99.5 | **<0.01** |
| | Late-onset-increasing | 2 | 22.1 | **<0.01** |
| Symptomatic-chronic | Late-onset-increasing | 2 | 100.0 | **<0.01** |
| **6-month** | | | | |
| Resilient | Intermediate-stable | 2 | 405.8 | **<0.01** |
| | Symptomatic-chronic | 2 | 729.1 | **<0.01** |
| | Late-onset-increasing | 2 | 63.9 | **<0.01** |
| Intermediate-stable | Symptomatic-chronic | 2 | 68.3 | **<0.01** |
| | Late-onset-increasing | 2 | 12.5 | **<0.01** |
| Symptomatic-chronic | Late-onset-increasing | 2 | 67.1 | **<0.01** |
| **1-year** | | | | |
| Resilient | Intermediate-stable | 2 | 344.3 | **<0.01** |
| | Symptomatic-chronic | 2 | 913.5 | **<0.01** |
| | Late-onset-increasing | 2 | 89.1 | **<0.01** |
| Intermediate-stable | Symptomatic-chronic | 2 | 173.6 | **<0.01** |
| | Late-onset-increasing | 2 | 5.5 | 0.06 |
| Symptomatic-chronic | Late-onset-increasing | 2 | 72.6 | **<0.01** |
| **2-year** | | | | |
| Resilient | Intermediate-stable | 2 | 358.5 | **<0.01** |
| | Symptomatic-chronic | 2 | 596.1 | **<0.01** |
| | Late-onset-increasing | 2 | 54.3 | **<0.01** |
| Intermediate-stable | Symptomatic-chronic | 2 | 55.8 | **<0.01** |
| | Late-onset-increasing | 2 | 17.2 | **<0.01** |
| Symptomatic-chronic | Late-onset-increasing | 2 | 39.3 | **<0.01** |
| **10-year** | | | | |
| Resilient | Intermediate-stable | 2 | 235.3 | **<0.01** |
| | Symptomatic-chronic | 2 | 772.6 | **<0.01** |
| | Late-onset-increasing | 2 | 417.2 | **<0.01** |
| Intermediate-stable | Symptomatic-chronic | 2 | 132.4 | **<0.01** |
| | Late-onset-increasing | 2 | 53.1 | **<0.01** |
| Symptomatic-chronic | Late-onset-increasing | 2 | 9.7 | **<0.01** |

*Bold indicates significance ($p$ < .05).*

*Abbreviations: df, degrees of freedom; LL, Log-likelihood.*

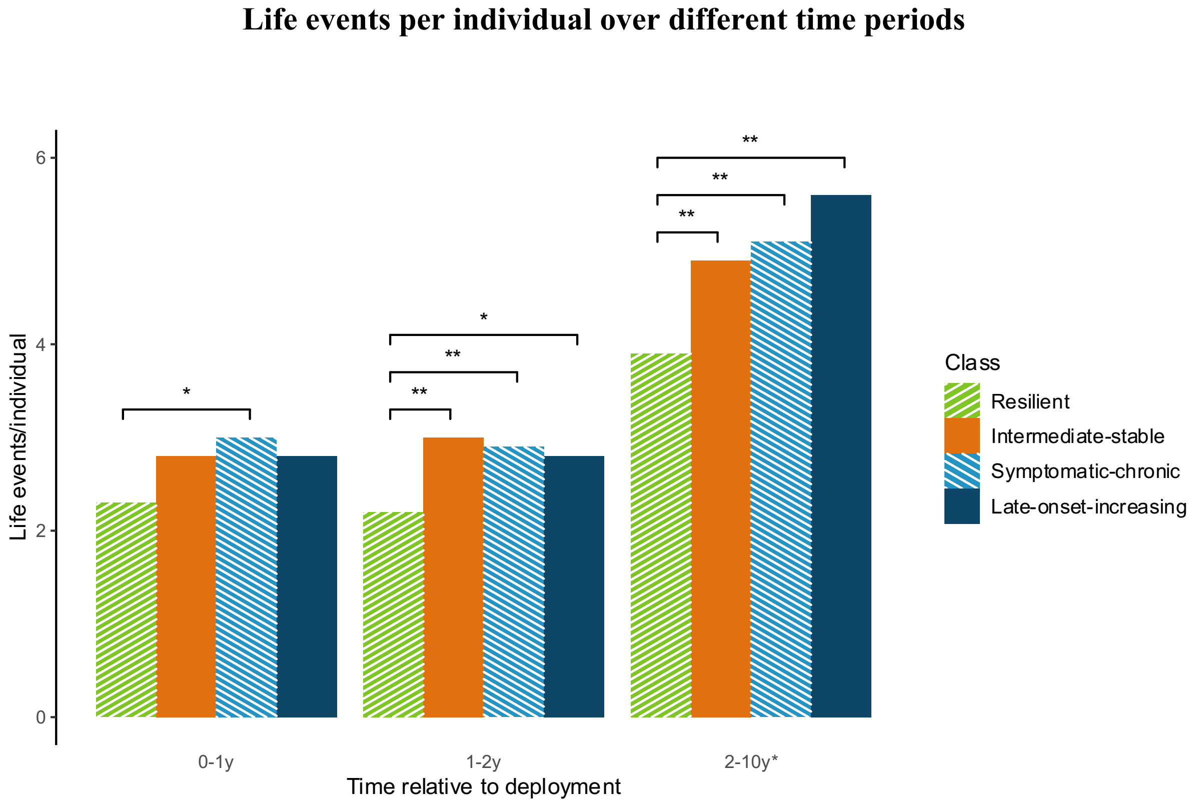


***Supplementary figure S11*** *– Figure to illustrate the amount of life events per individual over different time periods. Colors indicate different classes. Significant differences are presented with an asterisk ($* p < .05$, $** p < .01$).*

**Important life events were collected at the 5-year and 10-year time point. Participants reported life events in the past 6 months. For these comparisons, the results of the 5- and 10-year measurement are combined.*

**Supplementary table S12**

$p$-values of auxiliary models to assess the association between important life events and depressive symptoms at the 1-year time point.

| **Comparison** | | **1** | **2** | **3** | **4** | **5** | **6** | **7** | **8** | **9** | **10** | **11** | **12** | **13** | **14** | **15** | **16** | **17** | **18** | **19** | **20** |
|---|---|---|---|---|---|---|---|---|---|---|---|---|---|---|---|---|---|---|---|---|---|
| **Resilient** | **Intermediate-stable** | 0.80 | 0.48 | 0.31 | **<0.01** | 0.43 | 1.00 | 0.27 | 0.78 | 0.08 | 0.86 | 0.06 | 0.35 | 0.90 | 0.15 | 0.43 | 0.25 | **<0.01** | **<0.01** | 0.99 | 0.56 |
| | **Symptomatic-chronic** | 0.85 | 0.94 | 0.69 | 0.17 | 0.74 | **0.04** | 0.11 | 0.99 | **0.02** | 0.73 | **0.03** | **0.02** | 0.68 | 0.68 | 0.27 | 0.75 | **0.03** | **<0.01** | **<0.01** | 0.77 |
| | **Late-onset-increasing** | 0.49 | 0.17 | 0.55 | 0.81 | 0.70 | 0.80 | 0.55 | 1.00 | 0.90 | 0.58 | **0.03** | 0.42 | 0.94 | 0.52 | 0.28 | 0.21 | 0.06 | 0.42 | 0.10 | 0.34 |
| **Intermediate-stable** | **Symptomatic-chronic** | 0.73 | 1.00 | 0.71 | 0.39 | 1.00 | 0.06 | 0.11 | 1.00 | 0.47 | 0.87 | **0.00** | 0.18 | 0.78 | 0.85 | 0.17 | 0.61 | 0.34 | 0.51 | **0.02** | 0.88 |
| | **Late-onset-increasing** | 0.63 | 0.16 | 0.27 | 0.10 | 0.97 | 0.83 | 1.00 | 1.00 | 0.43 | 0.54 | **0.01** | 0.82 | 0.89 | 1.00 | 0.20 | 0.63 | 0.43 | 0.43 | 0.14 | 0.59 |
| **Symptomatic-chronic** | **Late-onset-increasing** | 0.48 | 0.17 | 0.44 | 0.32 | 0.96 | 0.08 | 0.58 | 1.00 | 0.22 | 0.49 | 1.00 | 0.46 | 0.74 | 0.84 | 1.00 | 0.41 | 0.94 | 0.24 | 0.72 | 0.54 |

*Bold indicates significance ($p < .05$).*

*Events: 1) failed an exam, 2) marriage, 3) new job, 4) conflict with employer, 5) move, 6) birth, 7) divorce/end of relationship, 8) retirement, 9) experienced an accident yourself, 10) accident of someone close, 11) being sick yourself, 12) sickness of someone close, 13) death of someone close, 14) burglary, 15) fire, 16) in touch with police, 17) relationship problems, 18) sexual problems, 19) financial difficulty, 20) change of household composition.*

### Supplementary table S13

$p$-values of auxiliary models to assess the association between important life events and depressive symptoms at the 2-year time point.

| **Comparison** | | **1** | **2** | **3** | **4** | **5** | **6** | **7** | **8** | **9** | **10** | **11** | **12** | **13** | **14** | **15** | **16** | **17** | **18** | **19** | **20** |
|---|---|---|---|---|---|---|---|---|---|---|---|---|---|---|---|---|---|---|---|---|---|
| **Resilient** | **Intermediate-stable** | 0.58 | 0.53 | 0.32 | **0.01** | 0.18 | 0.96 | **<0.01** | 0.70 | **0.02** | 0.84 | **0.04** | 0.65 | 0.12 | 0.96 | 0.21 | 0.84 | **<0.01** | **<0.01** | 0.41 | **0.03** |
| | **Symptomatic-chronic** | 0.07 | 0.23 | 0.80 | 0.09 | 0.78 | 0.07 | 0.83 | 0.34 | **0.01** | 0.18 | 0.51 | **0.01** | 0.49 | 0.47 | 0.10 | 0.68 | 0.05 | 0**<0.01** | 0.10 | 0.98 |
| | **Late-onset-increasing** | 0.81 | 0.21 | 0.09 | **<0.01** | 0.44 | 0.86 | 0.21 | 0.17 | 0.88 | 0.66 | **0.02** | 0.40 | 0.16 | 0.92 | 0.17 | 0.26 | 0.38 | 0.81 | 0.30 | 0.31 |
| **Intermediate-stable** | **Symptomatic-chronic** | 0.23 | 0.49 | 0.58 | **<0.01** | 0.53 | 0.13 | 0.05 | 0.58 | 0.42 | 0.20 | 0.47 | **0.01** | 0.11 | 0.50 | 0.05 | 0.62 | 0.28 | 0.17 | 0.39 | 0.18 |
| | **Late-onset-increasing** | 0.61 | 0.40 | **0.04** | **<0.01** | 0.16 | 0.89 | 0.48 | 0.33 | 0.21 | 0.61 | **0.01** | 0.31 | 0.67 | 0.90 | 0.62 | 0.37 | 0.14 | 0.10 | 0.64 | **0.04** |
| **Symptomatic-chronic** | **Late-onset-increasing** | 0.19 | 0.80 | 0.32 | **<0.01** | 0.41 | 0.32 | 0.39 | 0.67 | 0.09 | 0.62 | **0.01** | 0.34 | 0.11 | 0.69 | **0.04** | 0.24 | 0.58 | **0.01** | 0.83 | 0.38 |

*Bold indicates significance ($p < .05$).*

*Events: 1) failed an exam, 2) marriage, 3) new job, 4) conflict with employer, 5) move, 6) birth, 7) divorce/end of relationship, 8) retirement, 9) experienced an accident yourself, 10) accident of someone close, 11) being sick yourself, 12) sickness of someone close, 13) death of someone close, 14) burglary, 15) fire, 16) in touch with police, 17) relationship problems, 18) sexual problems, 19) financial difficulty, 20) change of household composition.*

### Supplementary table S14

$p$-values of auxiliary models to assess the association between important life events and depressive symptoms at the 10-year* time point.

| **Comparison** | | **1** | **2** | **3** | **4** | **5** | **6** | **7** | **8** | **9** | **10** | **11** | **12** | **13** | **14** | **15** | **16** | **17** | **18** | **19** | **20** |
|---|---|---|---|---|---|---|---|---|---|---|---|---|---|---|---|---|---|---|---|---|---|
| **Resilient** | **Intermediate-stable** | 0.45 | 0.70 | 0.33 | **<0.01** | 0.80 | 0.18 | 0.09 | 0.64 | 0.06 | 0.07 | 0.92 | 0.41 | 0.26 | 0.23 | 0.22 | **0.04** | **0.01** | **0.01** | **0.02** | 0.05 |
| | **Symptomatic-chronic** | **<0.01** | 0.33 | 0.50 | 0.10 | 0.48 | 0.23 | 0.07 | 0.94 | **0.02** | **0.01** | **0.02** | 0.23 | 0.50 | **0.04** | **0.01** | 0.12 | **<0.01** | **<0.01** | 0.14 | 0.37 |
| | **Late-onset-increasing** | 0.64 | **0.02** | 0.22 | **<0.01** | 0.96 | 0.48 | **<0.01** | 0.70 | 0.88 | 0.17 | 1.00 | 0.45 | 0.57 | 0.16 | 0.07 | 0.09 | **<0.01** | **<0.01** | **<0.01** | 0.05 |
| **Intermediate-stable** | **Symptomatic-chronic** | **0.02** | 0.27 | 0.97 | 0.53 | 0.43 | 0.06 | 0.64 | 0.71 | 0.41 | 0.38 | 0.06 | 0.11 | 0.88 | 0.34 | 0.20 | 0.99 | 0.35 | 0.10 | 0.79 | 0.63 |
| | **Late-onset-increasing** | 0.39 | **0.02** | 0.59 | 0.50 | 0.84 | 0.87 | **0.04** | 0.94 | 0.23 | 0.90 | 0.95 | 0.24 | 0.87 | 0.61 | **0.04** | 0.78 | **<0.01** | 0.41 | **0.01** | 0.56 |
| **Symptomatic-chronic** | **Late-onset-increasing** | **0.01** | 0.20 | 0.61 | 0.27 | 0.65 | 0.16 | 0.15 | 0.72 | 0.09 | 0.55 | 0.14 | 0.79 | 0.98 | 0.76 | **0.01** | 0.81 | 0.05 | 0.56 | **0.01** | 0.37 |

*Bold indicates significance ($p < .05$).*

*Events: 1) failed an exam, 2) marriage, 3) new job, 4) conflict with employer, 5) move, 6) birth, 7) divorce/end of relationship, 8) retirement, 9) experienced an accident yourself, 10) accident of someone close, 11) being sick yourself, 12) sickness of someone close, 13) death of someone close, 14) burglary, 15) fire, 16) in touch with police, 17) relationship problems, 18) sexual problems, 19) financial difficulty, 20) change of household composition.*

**Important life events were collected at the 5-year and 10-year time point. Participants reported life events in the past 6 months . For these comparisons, the results of the 5- and 10-year measurement are combined.*
